# Heating On Transport WitH prEtErm and Low birth weight infants using servocontrol thermoregulation (HOTWHEELS): study protocol for a randomised controlled trial

**DOI:** 10.64898/2026.09.29.26364248

**Authors:** Varunashree Chaudhary, Tess Watson, Michael Lee, Nicole Ilagan, Gracie Hodgson, Alex Wilson, Saumil Desai, Vamsi Batta, Jacqueline Gardiner, Matthew Cooper, Jonathan Davis

## Abstract

**Background:** Temperature instability during neonatal transport contributes to adverse outcomes in preterm and low birth weight infants. Hypothermia in this population is associated with increased risk of intraventricular haemorrhage (IVH), sepsis, and death. Current transport temperature management is reactive and manually controlled. The **Heating On Transport WitH prEtErm and Low birth weight neonates using Servocontrol patient temperature management system (HOTWHEELS)** trial aims to determine whether automated, servo-controlled thermoregulation provides superior thermal stability compared to standard care and reduces hypothermia-associated adverse outcomes.

**Methods:** This prospective, randomised, superiority trial is conducted by the Newborn Emergency Transport Service of Western Australia. Infants with gestational age of ≤34 weeks and/or weight of ≤1500 g requiring transport are randomised to servocontrolled or standard thermal care. The primary outcome is the proportion of infants with a temperature between 36.5–37.5 °C on arrival at the receiving neonatal unit. Secondary outcomes include moderate (<36.0°C) and severe (<35.0°C) hypothermia ≥15 minutes and temperature change during transport, IVH≤7 days, and mortality before discharge. Trial registration: ANZCTR12624000503527.

**Impact:** HOTWHEELS is the first randomised trial in neonatal transport to evaluate servo-controlled temperature control. We believe the findings will inform standards of temperature control during transport for preterm and low birth weight infants.

**Category of study:** Study protocol; special article

**Impact Statement:**

- Thermoregulation is an independent predictor of morbidity and mortality in neonates.
- Preterm and low birth weight infants are highly vulnerable to temperature changes due to immature physiological mechanisms.
- Achieving and maintaining normothermia is challenging within hospital settings, and even more so in neonatal transport environment.
- Bundled thermoregulation care still fails to achieve normothermia in over >20% transported neonates

## Introduction {9a}

Newborn infants regulate their body temperature poorly. Core temperature between 36.5°C and 37.5°C immediately after birth is essential for neonatal survival and avoidance of adverse outcomes (1,3,4). Low body temperature (<36.5°C) is common in infants born at hospitals, at home, and even in tropical environments (1). Preterm infants or those with very low birth weight (<1500g) are especially at risk of hypothermia due to their large surface area-to-body mass ratio, limited subcutaneous fat stores, reduced muscle tone, and limited ability to generate heat from fat reserves. In these infants, low admission temperature is strongly associated with mortality, with each 1°C decrease corresponding to a 28% increase in death rates (5). Moderate hypothermia has also been linked as a risk factor for intraventricular haemorrhage (6)

### Thermoregulation in Neonatal Intensive Care

The International Liaison Committee on Resuscitation (ILCOR) Neonatal Task Force has recommended maintaining the temperature of non-asphyxiated neonates between 36.5°C and 37.5°C from birth through admission and stabilisation. (7). In preterm infants <33 weeks’ gestation, admission temperature between 36.5°C and 37.2°C is associated with the lowest incidences of adverse outcomes, including severe neurological injury (grade III, IV IVH and periventricular leukomalacia), necrotising enterocolitis, bronchopulmonary dysplasia, and nosocomial sepsis (9). Temperature control is achieved through strategies that minimise heat loss via evaporation, conduction, convection, and radiation, and through active warming including maintaining adequate environmental temperature, overhead infant warmers, polyethene wraps, pre-warmed mattresses, and heated humidified respiratory gases (3). Despite these measures, hypothermia remains common at NICU admission (5,8,9,13,14). Thermoregulation bundles (adjuncts in combination) have consistently demonstrated reduced hypothermia and improved clinical outcomes in preterm infants ((15–18)

Automated thermoregulation systems (servo-controlled) use feedback from measured core temperature to detect and correct temperature deviations, enabling precise temperature control without continuous manual intervention. Their use has been reported in stabilising preterm infants after delivery and during interhospital transport (18–20), but optimal effectiveness is unclear (20).

### Temperature Control During Transport

Hypothermia during neonatal transport is a clinically significant adverse event. Incidence varies widely according to population characteristics, transport infrastructure, and the presence of specialised retrieval services. Up to 70% of transported infants are hypothermic at some point, and there is greater risk in infants with lower birth weight and gestation ((24) Environmental transitions, infant handling during stabilisation and transfer, limited humidity and radiant heat within transport environments, and unavoidable interruptions to thermal support all contribute to thermal instability, particularly in infants with immature thermoregulatory capacity (25)Hyperthermia may occur and has been reported in up to 19% of transported infants at admission. Infants experiencing moderate hypothermia (32°C to 36°C) experience the greatest mortality risk. Mortality appears improve when transport is facilitated by a specialised transport team (,24,25,26).

During transport, several devices are utilised to provide heat and prevent its loss. Transport incubators *(such as in* Figure 1A and 2) attached to a movable stretcher (Figure 2) are the mainstay of temperature control. These provide a regulated internal temperature set by the healthcare provider. Other adjuncts that provide heat include a thermal chemical mattress (e.g., TransWarmer®; Cooper surgical, CT, USA, Figure 1B). To prevent heat loss, sterile polyethylene cover, suit, or wraps (e.g., Neohelp Vygon®, Ecouen, France, Figure 1C) or hats and covers are used. A single study of a bundled approach during inter-facility neonatal transport reduced admission hypothermia from 52% to 17% without increasing hyperthermia or transport time (25).

**Figure 1.**
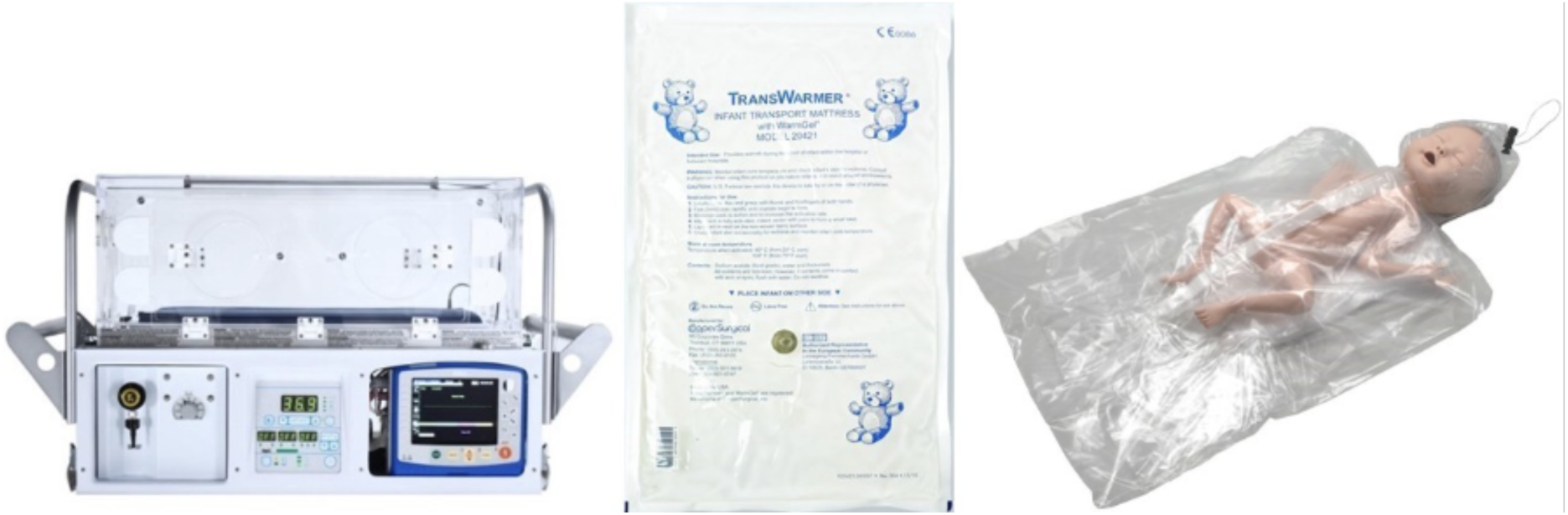
A: Airborne ® Voyager transport incubator courtesy of International Biomedical); B: Transwarmer chemical mattress; C: Neohelp Polythene Bag.

**Figure 2.**
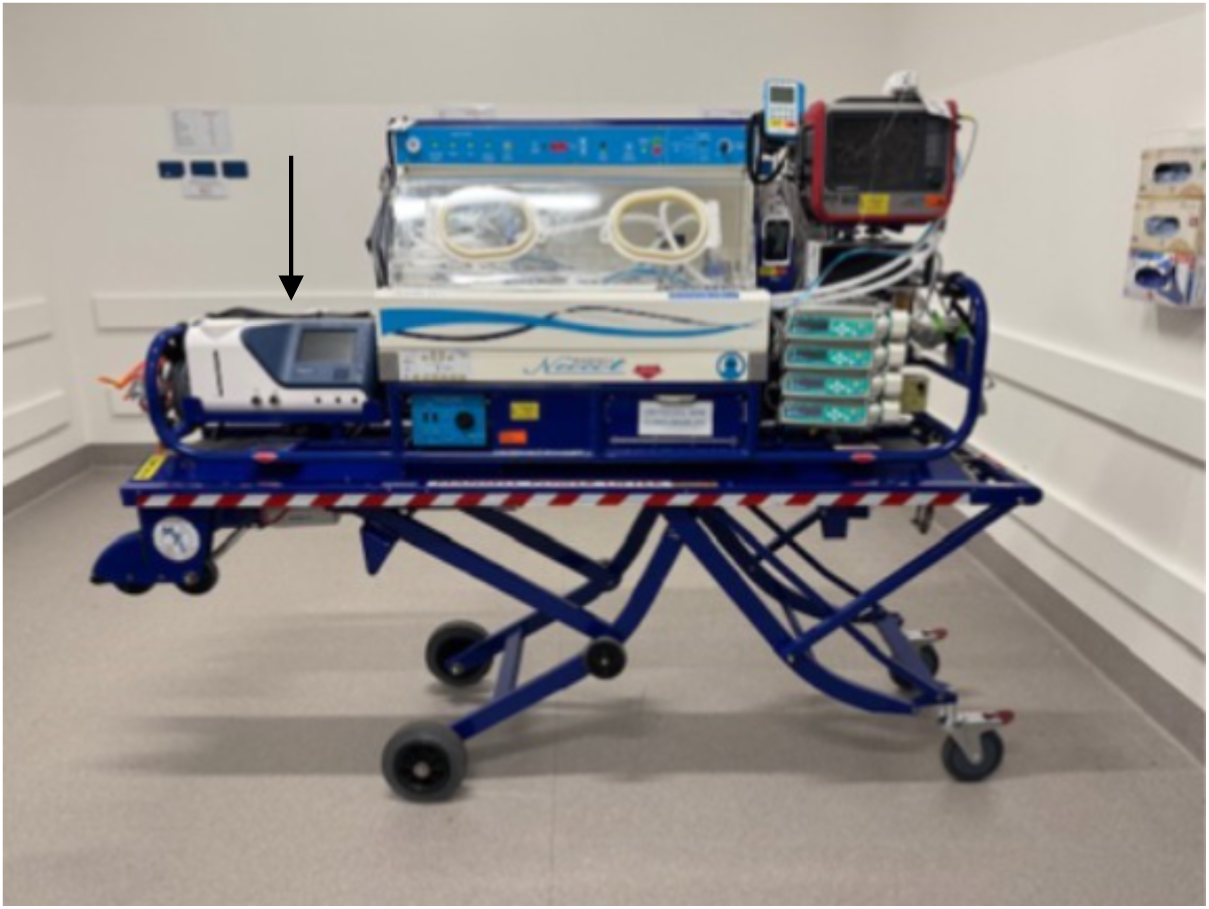
Criticool Mini mounted within a neonatal transport system. The Criticool Mini is indicated by the arrow.

Servocontrolled temperature management during transport has been reported in 36 extremely preterm infants, with the use of rectal core temperature monitoring. This observational study showed core temperatures increased from the beginning to the end of transport without complications ((20). No other studies of servo-controlled devices in transport exist. Further study of the device’s safety and efficacy during the transfer of preterm and low birth weight babies is necessary.

### Aims and Objectives {9b, 10}

This study aims to compare the effectiveness of a servo-controlled temperature management system with the current standard of thermal care in preterm and low birth weight infants during interhospital transport. We hypothesise that the servo-controlled device will achieve normothermia (36.5 to 37.5 C) on admission in a greater proportion of infants and reduce temperature fluctuations during transport. We further expect that the incidence of intraventricular haemorrhage will be lower in the servo-controlled group compared with standard care.

## Methods

### Participants, Interventions and Outcomes

#### Study Design and Setting {11, 12, 13}

The study is a single-centre, non-blinded, randomised controlled, superiority trial of thermal management, with a battery-enabled servocontrolled device (Criticool Mini, Belmont Medical Technologies MA, USA) against current ‘standard care’ (described below).

It will be conducted within the Newborn Emergency Transport Service of Western Australia (NETS WA). Infants can be referred for transfer from any healthcare facility in Western Australia to access specialised care not available at the referring site.

Infants requiring transfer <200 km are transported by road ambulance, while those requiring transfer >200 km are transported by aircraft to one of the two tertiary centres in Perth (the state capital): King Edward Memorial Hospital (KEMH) or Perth Children’s Hospital (PCH). Air transport may be via fixed or rotary-wing (helicopter).

For fixed-wing transfers, infants are transported by road ambulance from the referring hospital to the nearest airstrip, flown to Perth, and then transferred by road ambulance to the receiving NICU. Infants transferred by rotary wing have the same journey except they land on a helipad at PCH. If the end destination is King Edward Hospital a road ambulance transfer is required from PCH to KEMH. Infants remain in the transport cot for the entire duration of the journey until handover to the receiving NICU team. The NETS WA transport system has been described in detail previously (27).

Study design and protocol was reviewed by the Preterm Consumer Reference Group of the Kids Research Institute Australia (formerly Telethon Kids Institute) who provided specific input especially on the approach to parental consent in the immediate postnatal period. They will also contribute to the interpretation and dissemination of study findings through co-development of lay summaries and community updates.

#### Eligibility Criteria {14a}

All patients ≤34 weeks’ gestation and/or ≤1500g referred to the NETS WA for transport will be eligible for screening.

**Inclusion criteria:**

- Corrected gestational age ≤ 34 weeks and/or
- Current weight ≤ 1500g
- Informed consent

**Exclusion criteria:**

- Major congenital gastrointestinal abnormality
- Necrotising enterocolitis
- Retrieval situation where it is not possible for parent(s) to provide verbal consent.
- No informed consent

#### Explanation for the Choice of Comparators {9b}

The comparator was selected to reflect current neonatal transport thermal management practice within NETS WA and to ensure clinical relative and ethical equipoise. In routine practice, preterm infants requiring transport are managed using a bundled thermoregulation approach, incorporating transport incubator temperature control and adjunctive measures to minimise heat loss. These interventions are applied collectively rather than in isolation and represent the established standard of care for neonatal transport in Western Australia and many other transport services around the world.

The intervention was chosen to evaluate whether a battery-enabled, servo-controlled patient temperature management system provides incremental benefit compared with the established bundled approach. In managing patients with hypoxic ischaemic encephalopathy, servo-controlled devices are effective in attaining accurate target temperatures (28). There is little data on the benefit of closed-looped servo-controlled systems in neonatal intensive care and transport settings for preterm infants. Comparing the intervention against the current NETS WA practice allows assessment of clinical effectiveness, safety, and feasibility while maintaining alignment with routine care and preserving ethical equipoise, specifically in the transport environment.

#### Intervention Description {14b, 15a}

The NETS WA team will be dispatched with the capacity to provide both standard thermal care and servo-controlled temperature management. Following consent and randomised group allocation, the intervention will be applied, as soon as feasible, until admission to the NICU. All infants in the study will be transported in a portable incubator and neonatal transport system (Mansell, Toowoomba, QLD) with a mounted Servocontrolled temperature device Criticool Mini (Belmont Technologies MA, USA) (Figure 2).

##### Standard care

Infants randomised to the standard arm will be placed in the Airborne^®^ Voyager transport incubator (International Biomedical, TX, USA, Figure 1A) set to 33°C to 37°C. A TransWarmer^®^ thermal mattress (Figure 1B) is used as an additional heat source if the infant’s temperature is <36°C despite other corrective measures. The third element is a sterile polythene suit to minimise environmental heat loss (Neohelp Vygon, Figure 1C), used if the infant is less than 24 hours old, still requiring external humidity, or was born <27 weeks and is <2 weeks old. A hat is also placed on the baby’s head. The baby’s temperature is checked every 15 minutes by a skin probe readout or a manual axillary temperature measurement. The cot temperature is adjusted accordingly. The infant will receive this method of temperature management until admission to the receiving unit, where the final admission temperature will be recorded (Appendix 1).

##### Servocontrolled management

Infants randomised to the intervention arm will be managed using a battery-enabled servocontrolled device (Criticool mini, Belmont Technologies MA, USA) and infant wrap (Curewrap, Belmont Medical Technologies, MA, USA; Figure 3). The device is set to normothermia mode and a temperature of 37°C. It uses a rectal and skin thermometer. A 7G rectal thermometer (Medlinket, Shenzhen, China) is inserted 2cm per rectum. It is constructed from a smooth, medical-grade PVC sheath designed to allow atraumatic insertion and minimise the risk of mucosal injury. The incubator temperature is set at 35°C (or 33°C if baby is >5 days old). The infant is positioned in a wrap, a single-use, one-piece, body-shaped flexible garment. This is available in two weight-based sizes: one intended for babies weighing 2.5–4 kg, and another for babies weighing 4–7 kg. A nested configuration (Figure 4) is used to support developmental care in small infants. The infant will be placed in the nested wrap in preparation for transfer (Appendix 2).

**Figure 3.**
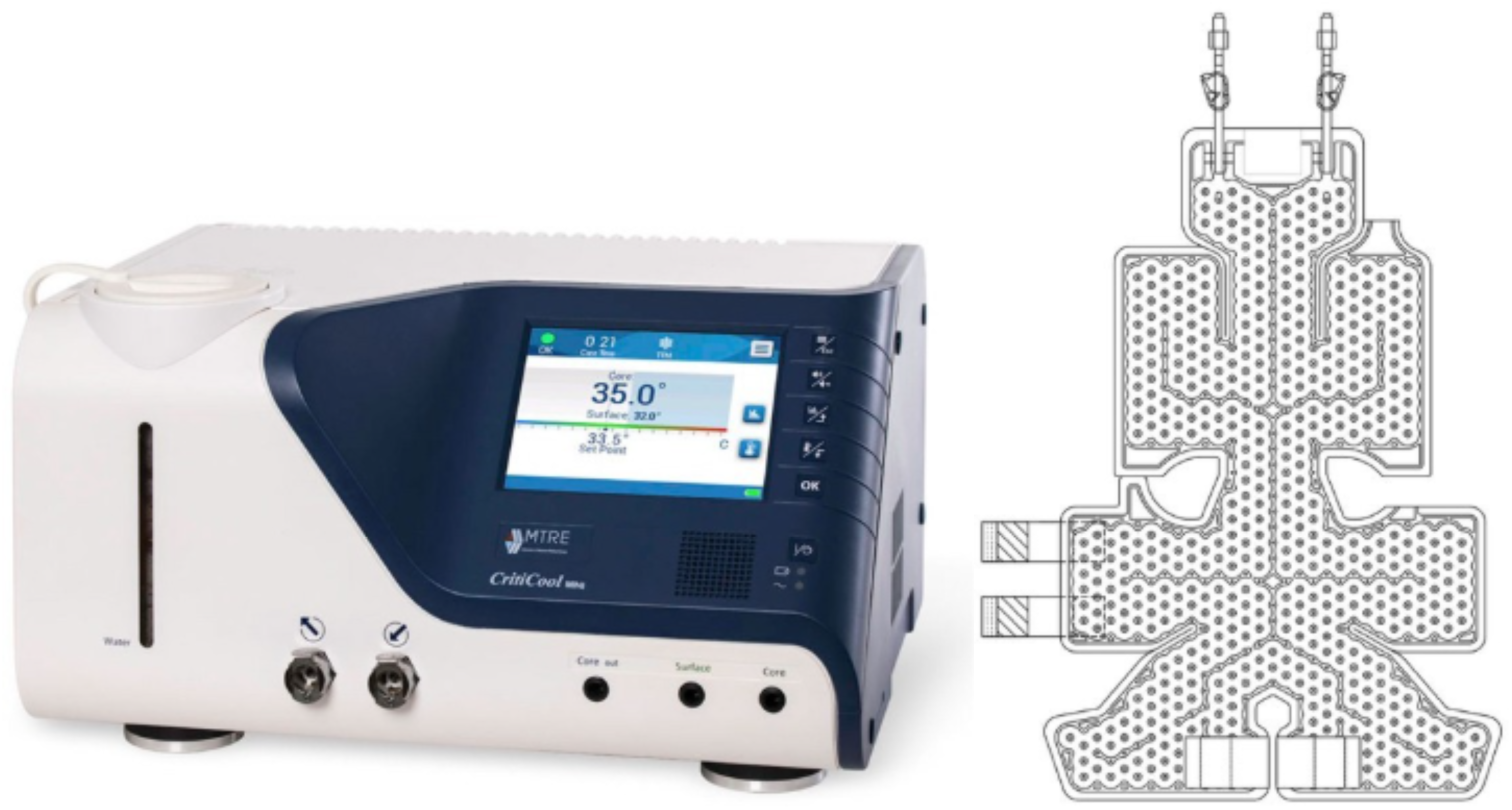
The battery servo-controlled device used was a Criticool Mini (Belmont Technologies MA, USA) and infant wrap (CureWrap, Belmont Medical Technologies, MA, USA).

**Figure 4.**
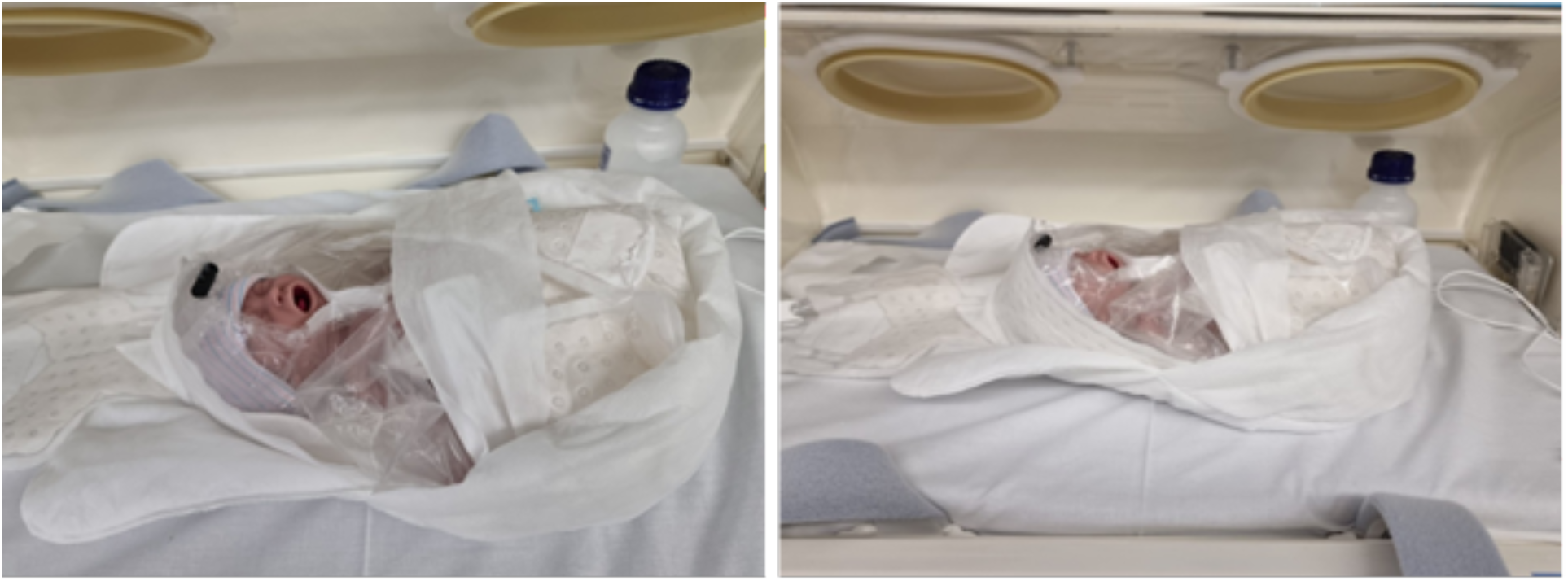
Adapted nesting of the plastic wrap for preterm infants.

To protect fragile skin, a BiliSoft (GE HealthCare, Waukesha, Wisconsin, USA) pad is placed between the infant and the wrap, Sil-Flex (Parker Healthcare, Victoria, Australia) tape is applied beneath the skin temperature probe sticker, and a Grip-Lok (TIDI Products LLC, Wisconsin, USA) device is utilised to secure the rectal probe to the thigh. A pressure relief algorithm periodically drains the water from the wrap for slight repositioning of the infant; specially designed channels within the garment distribute pressure. Careful positioning and nesting of the infant are used to support physiological stability and minimise focal pressure and shear forces. Positioning strategies aim to provide gentle containment and support, promoting flexed posture and even pressure distribution while avoiding focal areas of stress. These approaches are consistent with principles of developmentally supportive care aimed at reducing physiological stress in preterm infants (29,30,31).

Thermoregulation during transport is inherently dynamic and requires continuous monitoring. While the primary risk for preterm infants is hypothermia, overheating is also a possibility that must be promptly addressed. The ILCOR recommend that the temperature of newborns be maintained between 36.5°C and 37.5°C (inclusive), as both hypothermia and hyperthermia can increase the risk of neonatal mortality and morbidity (7). If the infant’s temperature rises above this target range, prompt stepwise adjustment of the temperature is required.

#### Criteria for Discontinuing or Modifying Allocated Interventions {15b}

Discontinuation of the allocated intervention will occur only in circumstances of catastrophic failure of the battery-enabled servo-controlled system or if there is a risk of extreme harm to the infant using the intervention. As clinical care may be compromised in this situation, the standard care method will be used instead. Non-adherence to the study protocol will be recorded, declared and described in any published manuscript of results, and analysis will be by intention-to-treat.

Major protocol deviations will include failure to apply the allocated intervention, significant interruptions to temperature management, or use of prohibited co-interventions. These will be documented and summarised, and sensitivity analyses (including per-protocol analysis) will assess their impact on study findings.

#### Strategies to Improve Adherence to Interventions {15c}

Intervention packs have been created as ‘grab bags’ included in a compartment beneath the cot. These will include patient recruitment materials, consent forms, temperature probes, the infant wraps, and action cards for a step-by-step guide for temperature management via both methods and data collection. The battery-powered, servo-controlled device is mounted on a transport system and carried on each transport. Staff training is recurrent and cyclical, and recruitment remains a regular educational focus. Staff are regularly trained on the device, including troubleshooting, and have access to educational videos. Staff also undertake regular simulations on recruitment and consent, to adapt to recognised best practice.

#### Relevant Concomitant Care Permitted or Prohibited During the Trial {15d}

Prior to randomisation, infants may have any local management deemed appropriate. This is not prescribed as part of the trial. Methods of thermal control and the temperature on arrival of the transport team will be documented on the patient case report form. Once randomised, infants in the intervention (servo-controlled arm) cannot have heating adjuncts added, such as a Transwarmer. Temperature management will be guided by a de-escalation/escalation protocol for each arm, which allows a dynamic response to persistent hypothermia or hyperthermia (Appendix 1 and 2).

#### Outcomes {16, 17}

##### Primary outcome

The primary outcome is the proportion of infants who are normothermic on admission to the receiving NICU, where normothermia is defined as an axillary temperature of 36.5°C to 37.5°C (inclusive). Temperature will be measured within 5 minutes of handover using a calibrated axillary thermometer by the transport team. The outcome will be analysed as a binary variable (normothermia vs non-normothermia).

##### Secondary outcomes

i. Short term

- The proportion of infants with mild, moderate and severe hypothermia (36.0 to 36.4°C) at time of admission;
- Proportion of infants with temperature >37.5°C (hyperthermia);
- Temperature at one hour of admission to the NICU;
- The change in temperature from first contact with the NETS team to arrival in NICU;
- Serious adverse events (SAEs) (hypothermia <35°C; hyperthermia >39°C).
ii. Safety considerations

- Ano-rectal, mucosal or skin trauma in relation to the rectal probe.
- Serious adverse events (SAEs) (hypothermia <35°C; hyperthermia >39°C).
iii. Prior to discharge

- The proportion in each group with intraventricular haemorrhage (all grades and grade III-IV, as measured on the Papile score (32) in the first 7 days of life;
- Mortality prior to discharge.

#### Sample Size {19}

The sample size calculation reflects the primary outcome of the proportion of infants with a temperature outside the normothermic range on arrival at the destination NICU – this is statistically equivalent to examining the proportion within the normothermic range. In NETS WA unpublished data from 2016– 2020, approximately 20% of infants (<34 weeks’ gestation and/or ≤1500 g) arrive outside the normal temperature range. We hypothesise that the use of servo-controlled thermal regulation will reduce this proportion to 5%, representing an absolute reduction of 15%. This effect size is supported by targeted care bundle interventions that have reduced admission hypothermia by 12% to >25% ((17,18,22,25,34) Achieving an arrival hypothermia rate of 5% or less is a meaningful clinical benchmark. In studies evaluating hypothermia in moderate to late preterm infants (<34 to 36 weeks’), establishing a 5% admission hypothermia threshold is safe and highly achievable in NICU, with post-intervention rates successfully dropping to as low as 2.6% to 4% (33,34). To detect this absolute difference of 15% (20% vs. 5%) using a two-sided comparison of proportions with a significance level (α) of 0.05 and over 80% power, 86 infants are required per group (172 total) recruiting 1:1. Calculations were performed using R (R Foundation for Statistical Computing, Vienna, Austria) (35)

#### Participant Timeline {18}

Potential participants will be identified when a neonate requires emergency transport and is referred to the NETS WA. Upon referral, the NETS WA clinical team will screen the infant for eligibility. A transport clinician will seek verbal informed consent from the parents or legal guardians. If granted, the infant will be randomised to receive either the study intervention or standard care, maintained throughout the transport journey. Data collection will conclude one hour after arrival at the cot side of the receiving NICU. It is mostly limited to routine clinical parameters. Axillary temperature will be recorded in the standard and intervention group at 15-minute intervals from first contact with the NETS team to one hour after arrival in NICU. In addition, the intervention group will have continuous rectal temperature monitoring which is a feature of the servocontrolled device. The primary study endpoint will be assessed at NICU admission. Additional outcome data including intraventricular haemorrhage (IVH) incidence and grade severity and mortality will be collected up to hospital discharge and is part of routine collection. Incidence and grade of IVH will be determined from the independent reports of routine medical imaging examinations on day 1 and 7 which use the Papile score for reference (32).

#### Recruitment {20}

Premature and low birth-weight infants (≤34 weeks and/o 1500g) who are referred to and transferred by NETS WA to King Edward Memorial Hospital (tertiary perinatal centre) or Perth Children’s Hospital (quaternary neonatal referral centre) will be screened for eligibility by the transport team at the time of referral. To promote adequate enrolment and maintain protocol awareness across the transport service, team will be provided with regular educational briefings and screening aids by NETS WA coordinators, alongside monthly reviews of transport logs to capture any missed eligible infants.

If eligible, both written and verbal information is provided, and consent is obtained verbally at the reference site. Written consent will be confirmed within 72 hours of arrival. This phased two-step approach has previously been used in clinical research within NETS WA (36) and is specifically utilised to reduce immediate psychological burden on families during the acute and stressful window of emergency neonatal transport. Recruitment will likely take place over a 3-year period.

### Assignment of interventions and allocation

#### Sequence Generation {21a, 21b}

Infants will be randomly assigned to intervention arm or standard thermal care (control arm) at a 1:1 allocation ratio. Multiple births will be randomised as individuals, as they are referred and transferred independently.

The allocation sequence will be computer-generated using the randomisation module within REDCap (37). To ensure strict concealment and minimise selection bias, the sequence will be generated and uploaded into REDCap by the trial statistician (MC) who is not involved in participant recruitment, clinical care, or day-to-day trial operations.

Randomisation will be restricted using permuted blocks (size 4 and 6). To ensure a balance across critical baseline prognostic factors, randomisation will be stratified according to two criteria:

- **Infant weight:** ≤1000g and >1000g.
- **Transport modality:** Air versus road transport.

Based on transport data collected between 2023 – 2026 we anticipate 40-45% of infants in the final cohort will be ≤1000g (n∼75) and 15 - 20% will transported by air (n∼25).

To prevent predictability of the allocation sequence, the specific block sizes and their random order within each stratum will be maintained in a separate document held by the statistician and will remain completely unavailable to the transport teams and clinical staff enrolling participants.

#### Allocation Concealment Mechanism {22}

To ensure strict allocation concealment, a centralised, web-based randomisation sequence will be implemented within the secure REDCap platform, completely hidden from the NETS WA transport team. Randomisation will only occur at the bedside after the infant’s eligibility criteria have been verified, baseline data are completed, and verbal consent is obtained.

Upon submitting these irreversible, mandatory pre-randomisation data into the secure REDCap web interface, the system will dynamically and automatically reveal the assigned treatment arm. This real-time, software-controlled mechanism ensures that the upcoming group assignment cannot be accessed, anticipated, or manipulated in advance by the personnel enrolling the participants.

#### Implementation {23}

Trial-trained neonatal transport doctors or the project’s research nurse will assess infant eligibility and enrol participants prior to travel. Upon enrolment and consent, these same authorised personnel will access the REDCap randomisation module to assign the infant to their respective intervention group. A detailed timeline of allocation, interventions, and assessments is illustrated in the SPIRIT diagram (Figure 5).

**Figure 5.**
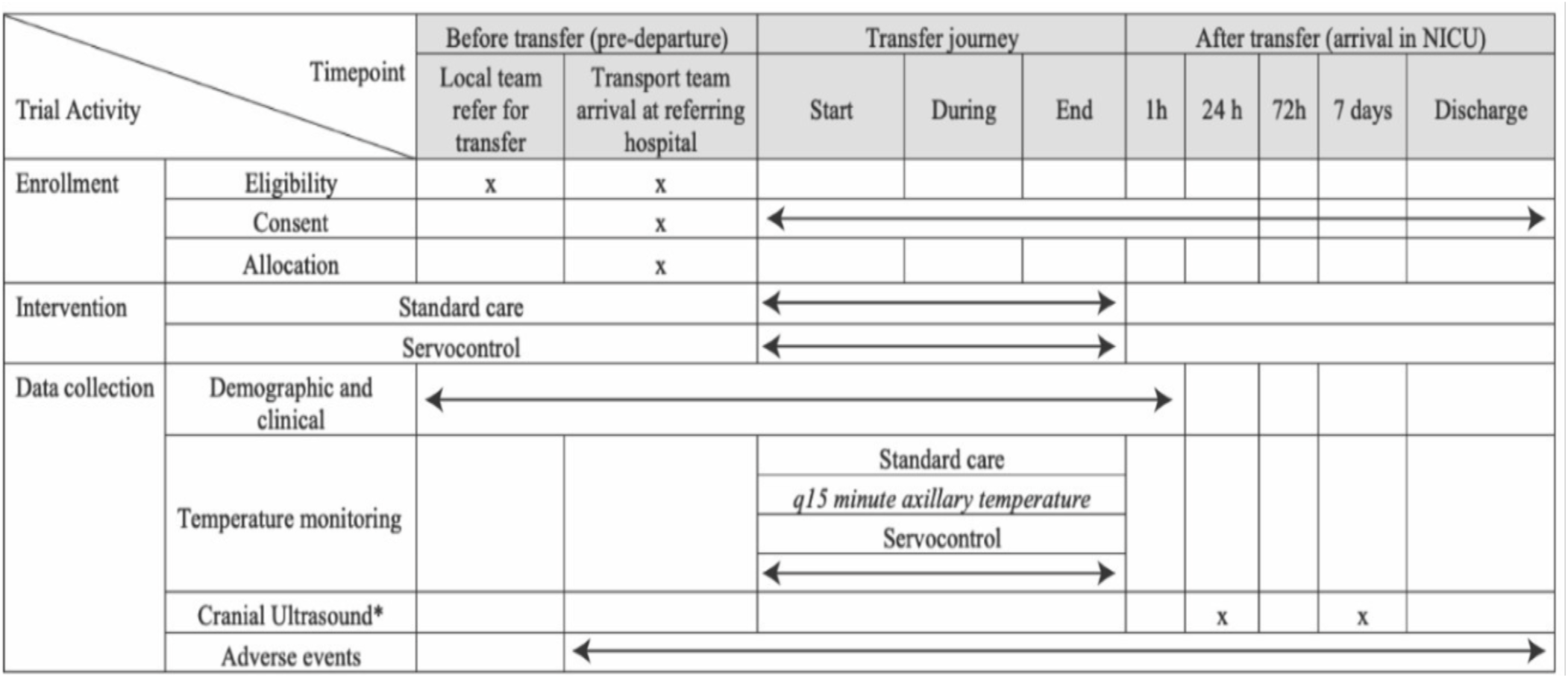
Schedule of enrolment, interventions, and assessments across pre-transfer, transport, and post-transfer phases (SPIRIT). Clinical and demographic data are collected from referral to completion of transport as part of routine processes. Adverse events are monitored from referral to discharge. “X” denotes discrete timepoints and arrows indicate continuous activities.

#### Who Will Be Blinded {24a}

Due to the physical nature of the intervention, blinding of the trial participants (parents) and the treating clinical team is not possible. To minimise measurement bias, temperature measurement at NICU admission will be performed using standardised unit procedures and calibrated devices, with staff trained in consistent measurement techniques.

The trial statistician conducting the analysis and the data managers will remain strictly blinded to treatment group allocations until the database is locked and the analysis plan is executed.

#### Procedure for Unblinding if Needed {24b, 24c}

As this is an open-label trial where the clinical team and participants are fully aware of group allocation, a formal procedure for emergency unblinding is not applicable.

### Data Collection, Management, and Analysis

#### Data Collection Methods {25a}

Data will be collected prospectively into the REDCap electronic transport database and the trial database. Throughout each transport, clinical staff will contemporaneously complete a case report form (CRF) in addition to the standard clinical data collection tool. Data will be entered directly into the REDCap database during the transport. For infants in the intervention arm, continuous temperature data (skin and rectal) are recorded via a skin sensor and rectal probe. Data are stored in a data logger (Clinilogger module, Belmont Medical Technologies, v1.6.3, MA, USA) attached to the servocontrolled temperature device. After the cot returns to base and the patient handover is complete, the Clinilogger is removed, and data are extracted as a CSV file.

#### Data Management {26}

A composite trial database will be established using REDCap, hosted by the Department of Health, Western Australia. Baseline clinical data from standard NETS WA transport collection will be ‘cross-piped’ with research trial data to form this single, unified composite database.

To ensure data integrity, the NETS transport research nurse will check all staff-entered data for quality and consistency. The REDCap database will utilise in-built range and consistency checks. Furthermore, a formalised data management plan will be followed, featuring monthly reviews and data verification by the research nurse to check for missing data or entry errors. Regular audits will be completed by research staff.

The REDCap database is password-protected, integrated with two-factor authentication, and restricted strictly to study-approved members of the NETS Team via WA Health-registered devices (iPads, tablets, computers, or laptops). Hard copies or digital copies of consent forms and CRFs will be saved in a dedicated, password-protected trial file on a secure WA Health server.

All data stored on the WA Health REDCap system is accessible only under WA Health security access protocols. No identifiable data will be utilised in the analysis or reporting of outcomes, ensuring no participant can be identified in any public output. The Chief Principal Investigator will have full access to the final dataset. De-identified data may be made available to external researchers upon reasonable request, subject to institutional governance and data sharing agreements.

#### Confidentiality {33}

To ensure participant confidentiality, the study database in REDCap will utilise a re-identification key system. While the primary database contains patient identifiers, REDCap automatically assigns a unique, non-identifying study ID number to each participant. Only de-identified, coded datasets will be extracted from REDCap for analysis by the study team and the biostatistician. Re-identification will only be possible by authorised study team members who possess specific administrative access rights to the master database.

All physical source documents and hard copies of data will be filed securely in a locked cabinet within a restricted-access office. Data analysis and reporting will be performed strictly by the approved study team and the designated study biostatistician, ensuring no participant is identifiable in any public output or publication.

At the conclusion of the trial, data will be archived and securely stored for a minimum of 25 years, in compliance with Section 12.4.4.8 of the Western Australian Sector Disposal Authority. Following the mandatory retention period, data destruction will be executed in accordance with Child and Adolescent Health Service (CAHS) policy, documented, and formally listed with the relevant disposal authority. Physical files will be shredded and disposed of via secure confidential waste bins, and all digital records will be permanently erased.

#### Statistical Analysis Plan {27}

Statistical analyses will be conducted on an intention-to-treat (ITT) basis, encompassing all randomised infants according to their initial group allocation, regardless of protocol adherence or subsequent deviations. Descriptive statistics for baseline demographic and clinical characteristics will be summarised by treatment group. Continuous variables will be reported as means with standard deviations or medians with interquartile ranges, depending on distribution, while categorical variables will be presented as counts and percentages. No formal statistical comparisons of baseline characteristics will be undertaken.

The primary outcome, defined as the proportion of infants achieving normothermia upon admission to the NICU, will be compared between treatment groups using logistic regression adjusted for the randomisation stratification variables. The primary treatment effect will be presented as an odds ratio (OR). In addition, absolute risk differences and risk ratios will be estimated where appropriate to aid clinical interpretation. Secondary outcomes will be evaluated using regression models tailored to their underlying distributions. Specifically, binary outcomes will be analysed using logistic regression and reported as ORs, continuous outcomes will be analysed using linear regression and reported as mean differences, ordinal outcomes will be analysed using ordinal logistic regression and reported as ORs, and count outcomes will be analysed using Poisson regression or negative binomial regression in the presence of overdispersion and reported as incidence risk ratios. If distributional assumptions are violated, appropriate data transformations or non-parametric alternatives will be explored. All statistical tests will be two-sided with a significance threshold of alpha = 0.05, with treatment effects reported with 95% confidence intervals.

To optimise the precision of the treatment effect estimates, both minimally adjusted (stratification variables only), and covariate adjusted analyses will be reported. Covariate adjusted models will control for prespecified, clinically relevant baseline covariates, including birth weight and distance travelled during transport. The extent and patterns of missing data will be systematically evaluated and described. If missing data is substantial (> 5%) and satisfies the missing at random assumption, multiple imputation by chained equations will be utilised.

To evaluate heterogeneity of the treatment effect, exploratory subgroup analyses will be performed by introducing an interaction term between the treatment group and the respective subgroup variable. Subgroups of interest include birth weight (≤1000 g vs >1000 g), transport modality (road vs air transport), and chronological age at referral (≤24 vs >24 h; to compare efficacy in the immediate postnatal transitional period given older chronological age my influence thermoregulatory stability and response to the intervention). All subgroup analyses are exploratory in nature, are not powered for definitive conclusions, and will be interpreted with caution.

Sensitivity analyses will be conducted to assess the robustness of the primary findings to deviations from model specifications and assumptions regarding missing-data mechanisms. As a supportive analysis, a per-protocol analysis will also be performed, which will exclude infants with major protocol violations to provide an efficacy-focused estimate of the intervention. All statistical analyses will be performed using R (R Foundation for Statistical Computing, Vienna, Austria) (35).

### Monitoring

#### Data Monitoring Committee {28a, 28b}

An independent Data and Safety Monitoring Committee (DSMC) will be established to safeguard the interests of trial participants and monitor safety data. The DSMC will operate externally and completely independent of the trial steering committee and sponsor, comprising a biostatistician and three clinical specialists with no competing conflicts of interest. The DSMC will have access to unblinded, comparative data and will review cumulative serious and non-serious adverse events according to the strict timeframes and procedures predefined in the formal DSMC Charter. This charter, outlining the committee’s precise roles, responsibilities, constitution, and operational guidelines, will be reviewed and approved by all members prior to the randomisation of the first participant.

No interim analyses for efficacy or futility will be conducted. Consequently, no formal statistical stopping boundaries are defined. The DSMC will monitor safety data continually, and any recommendation to terminate or modify the trial early will be based on clinical judgment regarding emergent safety concerns. The final decision to act upon a DSMC recommendation to cease the trial rests with the Chief Principal Investigator (CPI).

#### Adverse Event Reporting and Harms

All Serious Adverse Events (SAEs), safety-critical events, or urgent safety measures identified by an investigator will be reported to the Sponsor within 24 hours of awareness, in accordance with the NHMRC Safety monitoring and reporting in clinical trials involving therapeutic goods (2016) (38) guidelines. For any SAE assessed by the local Principal Investigator (PI) as potentially, probably, or related to the trial intervention, an expedited review will be undertaken by the Data and Safety Monitoring Committee (DSMC) according to the timeframes outlined in the DSMC Charter.

All standard, non-serious Adverse Events (AEs) will be recorded in the participant’s case report form and monitored by the Chief Principal Investigator (CPI). Any local clinical incidents arising from an AE will be documented and investigated in alignment with the institution’s clinical incident management system. Any Significant Safety Issues (SSIs) or Suspected Unexpected Serious Adverse Reactions (SUSARs) occurring at a study site will be reported to the participating institution’s Research Governance Officer and the CPI within 72 hours of the site PI becoming aware of the event.

#### Frequency and Plans for Auditing Trial Conduct {29}

Trial audits will be scheduled to occur dynamically after the enrolment of every 66 participants (representing sequential milestones throughout the study duration). These will be operationally synchronised with the scheduled data reviews of the independent Data and Safety Monitoring Committee (DSMC).

The audits will be conducted for quality assurance and will specifically evaluate the integrity of the informed consent process, the accuracy and timeliness of data entry into the case report forms, and strict protocol adherence to temperature management processes. Any deviations or quality issues identified during these audits will be documented, addressed through formal corrective and preventive actions (CAPA), and reported directly to the Chief Principal Investigator (CPI) and the Trial Steering Committee.

#### Ethics and Dissemination {8, 30}

Ethical approval for this trial has been obtained from the Child and Adolescent Health Service Human Research Ethics Committee (HREC; Reference: RGS0000006433) Trial-related monitoring, audits, and regulatory inspections will be permitted, granting direct access to source documents and medical records to authorised representatives from external sponsors, the HREC, and institutional governance review bodies.

The trial is registered with the Australian New Zealand Clinical Trials Registry (ANZCTR; Reference: ACTRN12624000503527). The findings of this study will be disseminated via publication in peer-reviewed scientific journals, presentations at national and international neonatal conferences, and through institutional reporting channels. No individual participant or clinical staff member will be identifiable in any dissemination output.

#### Plans for Communicating Protocol Amendments {31}

Protocol modifications will be kept at a minimum and only considered when clinically or operationally essential. All substantial amendments will be submitted to the CAHS HREC for review and approval.

Amendments will be communicated to the clinical teams and investigators, and via education sessions, daily briefs, simulation training and written communications. The clinical trial registry will be updated to reflect these revisions within (e.g., 30 days) of approval. The full study protocol will be made publicly available as part of the journal publication and/or as supplementary material. The protocol is also accessible via the trial registration record (ANZCTR). Any subsequent protocol amendments will be updated in the registry and reflected in the published version as appropriate.

#### Consent {32a, 32b}

Transport of an unwell infant is a distressing experience for families. To accommodate this, a two-step informed consent pathway will be used to reduce the potential emotional and cognitive burden on the family. This method has been successfully used in previous studies (36).

At the referring hospital, a clinician from the NETS WA team will provide a brief, verbal overview of the study to the available parents, including a summary of procedures, risks, and expected benefits. This will be supported by written information. Verbal consent to randomise the infant will be obtained and documented in the clinical notes. The parents will be given the Participant Information and Consent Form (PICF). The transport research nurse or transport clinician will approach the family within 72 hours of admission for a formal, written informed consent to confirm ongoing trial participation and data collection. Parents retain the right to withdraw their infant from the study at any stage without prejudice to their clinical care. No ancillary studies or secondary biobanking of specimens are planned as part of this trial, and no additional consent for secondary data use will be sought.

#### Dissemination Plans {11, 34}

Trial results will be disseminated to healthcare professionals, participants, and the broader public through multiple channels, with no restrictions on publication. The primary findings will be submitted to a peer-reviewed open-access journal, and abstracts will be presented at relevant national and international scientific conferences to facilitate academic discussion and knowledge translation.

To maximise community relevance and impact, consumer groups and parent representatives involved in the study’s funding and ethical approval processes will play an active role in dissemination. They will contribute to the co-development of plain-language summaries and community updates, ensuring that results are accessible and meaningful to families and the public. Trial outcomes and acquittal reports will also be provided to the funding body, the Perth Children’s Hospital Foundation, and other key stakeholders.

In line with open science principles, there are currently no plans for ancillary sub-studies using the trial data. However, following publication of the primary results, the de-identified participant-level dataset and statistical analysis plan will be securely archived. These materials will be made available for future research upon reasonable request to the corresponding author.

#### Ancillary and Post-Trial Care

Participants will receive all routine clinical care from NETS WA throughout their journey to the receiving hospital and NICU care in the respective unit, independent of trial participation. No additional post-trial interventions are required as the study intervention is limited to the transport period. Measurements pertinent to the trial, after transfer has concluded include an axillary temperature 60 minutes from admission, the day 1 and 7 cranial ultrasound and the date of discharge or death.

In the event of harm related to trial participation, clinical care will be provided according to standard institutional protocols. Indemnity, insurance, and compensation arrangements are covered under the governance framework of the CAHS and WA health system policies. Participants and their families will have access to appropriate clinical follow-up and support as required.

## Discussion

Maintaining normothermia is critical for optimal neonatal outcomes, particularly in the immediate postnatal period. Effective temperature regulation is especially important in preterm infants, who are highly vulnerable to thermal instability. Despite the use of adjunctive strategies to provide heat and minimise heat loss, hypothermia remains common during neonatal transport (4,20,24–26,39). Outborn infants represent a particularly vulnerable population. The *Lancet Child & Adolescent Health Commission on the Future of Neonatology* highlights the ongoing disparities in outcomes for infants requiring transfer and emphasises the importance of optimising systems of neonatal care. Improving the quality and safety of neonatal retrieval is therefore a key priority, with potential for meaningful impact on neonatal outcomes (24–26,39)

Servocontrolled systems provide an automated approach to thermal management and have the potential to improve temperature stability during transport. There is limited evidence regarding their safety and efficacy in the neonatal retrieval setting. There are very few prospective trials conducted in neonatal transport, and this study addresses an important evidence gap in a logistically challenging and under-researched clinical environment.

This study is pragmatic and embedded within an established neonatal retrieval service, focused on a high-risk outborn population, and tests a clinically relevant intervention in a real-world setting. It addresses a recognised gap in neonatal transport research, where high-quality prospective data are scarce.

## Data Availability

The manuscript is a trial protocol. No data is presented.

