## Appendix 1 for "Heating On Transport WitH prEtErm and Low birth weight infants using servocontrol thermoregulation (HOTWHEELS): study protocol for a randomised controlled trial"

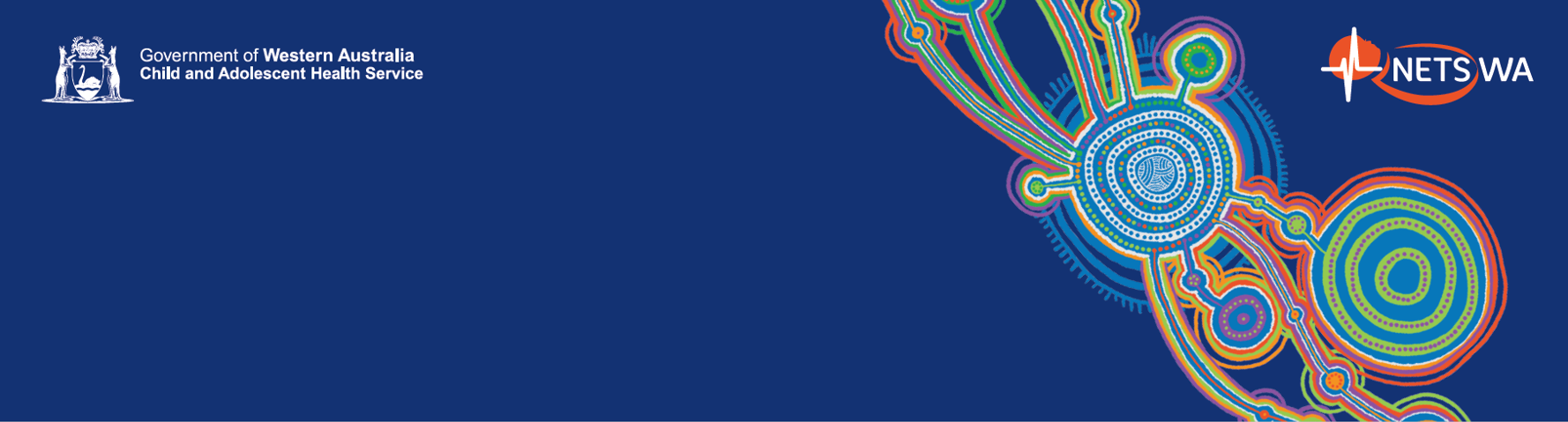
HOTWHEELS STANDARD TREATMENT ARM (A)

**Prior to arrival referring hospital**

Plug cot to power source in ambulance/plane

Set cot temp 33-37ºC* using **Diagram 1 (Appendix 3)**

Prewarm hat, linen and nest, stethoscope and Neohelp^TM^ inside cot

If needed switch on Neopod humidifier to 37-39ºC + prime with sterile water just prior to/on arrival

(Have chemical mattress^+^ placed in cot for baby < 25wk GA to lie on but **DO NOT** prime yet)

*Starting cot temp may vary depending on weather conditions e.g. if 40+°C weather, consider lower cot temperature. When using chemical mattress have white side up and pre-warmed linen between baby and mattress. Be careful when Voyager cot temp reaches more than 39°C – cot may alarm or turn off.

During transfer check all port holes and covers properly sealed to avoid drafts

Consider extra towels to cover cot (but keep baby still visible ) in windy/cold conditions

**On arrival**

Plug transport cot to power source and check hospital overhead heater turned ON

Check axilla temp, remove any wet clothing/towels (Consider using Servocontrol skin probe temp on Giraffe)

Place baby in Neohelp^TM^ if Place baby in Neohelp^TM^ if <24 hours old, baby is requiring external humidification, or was born <27 weeks and is <2 weeks old (**Diagram 2**), Bilinest and surround with warm towel

Apply skin probe temp and record **surface temp every 15 mins & axilla temp every 30 mins**

**Start filling in ‘HOTWHEELS STANDARD TREATMENT REDCAP FORM’**

Remove chemical mattress (if used)

Recheck temp every 15 mins

 Reduce cot temp by 0.5-1ºC

Apply chemical mattress (if not used)^+^

Increase cot temp by 0.5-1ºC

Recheck temp every 15 mins

**If temp >37.5**ºC

**If temp <36.5**ºC

**If temp <36.0**ºC

Increase cot temp by 0.5ºC

Recheck temp every 15 mins

**When temp between 36.5-37.5ºC**

Check lines and ventilation, hat (+ Neohelp) secured and assign team members roles to prepare for transfer

Consider adjusting cot temp to same temp as overhead heater temp if needed, otherwise use **Appendix 1** as a guide

Transfer baby under a warm blanket (in the Neohelp +/- chemical mattress if used) into cot

Reduce overhead heater temp by 25-30%/0.5-1ºC

Recheck axilla temp every 15 mins

Change heater to manual mode & adjust % if temp still >37.5ºC

**If temp >37.5ºC**

**If temp <36.5ºC**

**If temp <36.0֯C**

Apply chemical mattress^+^ & increase overhead heater temp 25-30%/0.5-1ºC

Recheck axilla temp every 15 mins

Change heater to manual mode & adjust % if temp still <36ºC

Increase overhead heater temp by 10-15%/0.5ºC

Recheck axilla temp every 15 mins
