## Appendix 2 for "Heating On Transport WitH prEtErm and Low birth weight infants using servocontrol thermoregulation (HOTWHEELS): study protocol for a randomised controlled trial"

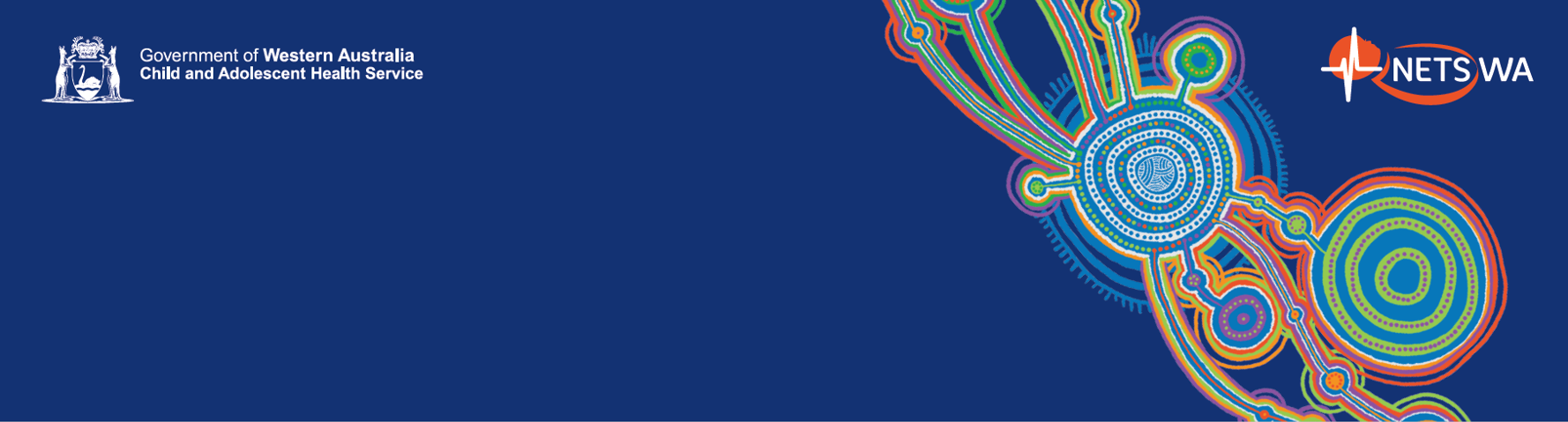

**Diagram 1— Cot Starting Temp Algorithm**

| <1000gms | 0-24hrs of life | 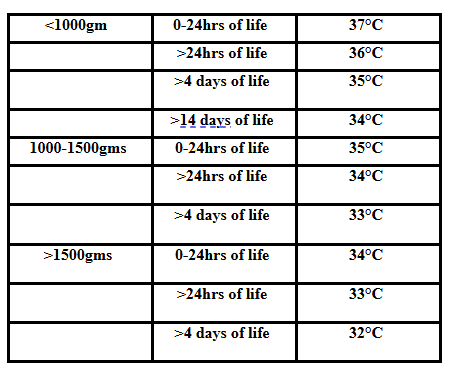37 ˚C |
| --- | --- | --- |
|  | >24 hours of life | 36 ˚C |
|  | >4 days of life | 35 ˚C |
|  | >14 days of life | 34 ˚C |
| 1000-1500gms | >0-24 hours of life | 35 ˚C |
|  | >24 hours of life | 34 ˚C |
|  | >4 days of life | 33 ˚C |
| >1500gms | >0-24 hours of life | 34 ˚C |
|  | >24 hours of life | 33 ˚C |
|  | >4 days of life | 32 ˚C |

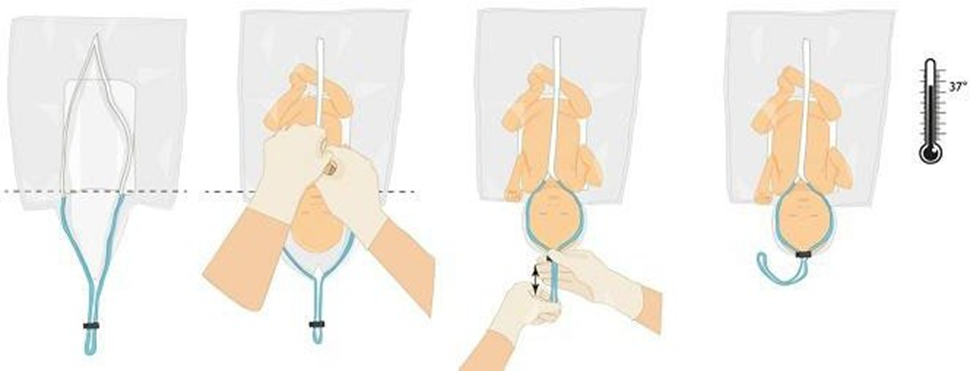

**Diagram 2—Neohelp Protocol**

Open Neohelp^TM^ , prepare a small hole for the right hand to apply oxygen saturation probe then place infant in the bag

Draw the string around the infant’s head and close the Velcro with the umbilical cord and clamp external to the bag.

Put a wool like hat on the infant’s head over the plastic Neohelp^TM^

Bring the right hand OUTSIDE the Neohelp^TM^ in the small hole that has been pre- made for placement of oxygen saturation probe.

Always transfer infant in a Neohelp^TM^ within a blanket to avoid slipping on transfer

If Neohelp^TM^ unavailable the NeoWrap^TM^ can be used with a knitted hat.

Prior to transfer measure the infant’s temperature and if not in the normal range address this prior to transfer.
