## Appendix 3 for "Heating On Transport WitH prEtErm and Low birth weight infants using servocontrol thermoregulation (HOTWHEELS): study protocol for a randomised controlled trial"

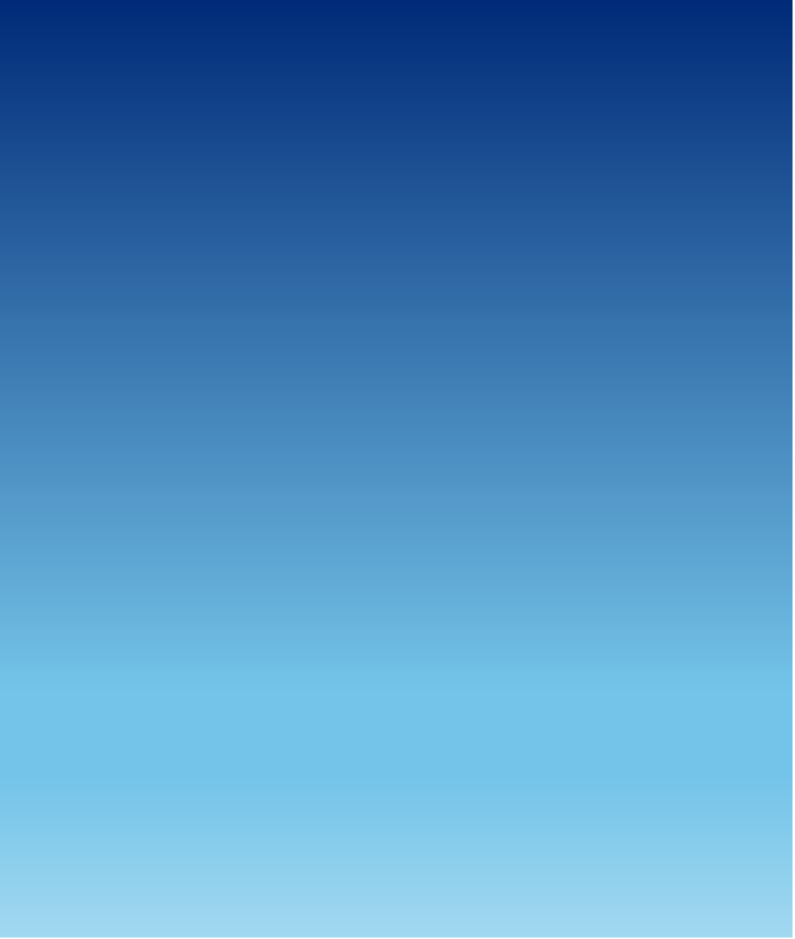

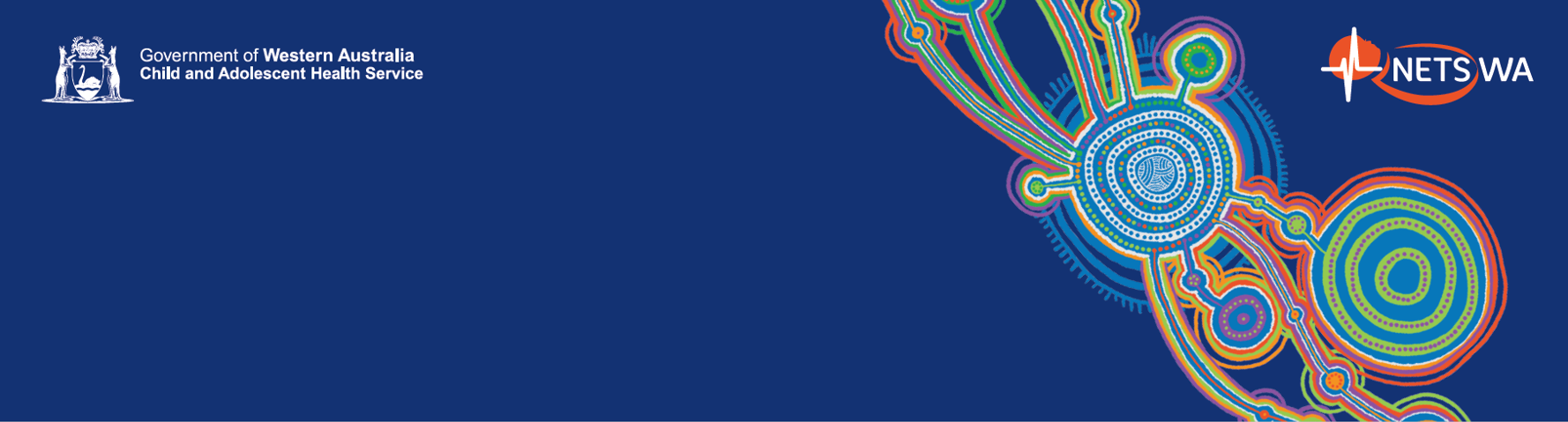
HOTWHEELS CRITICOOL TREATMENT ARM (B)

- Check baby regularly for skin pressure sores and document on body map
- Alarm of the Criticool may go off on transfer due to water flow obstruction - Check all hoses unkinked.
- Criticool device has only 1 hour of battery charge and should to be plugged in at all times.
- Check axilla temp and rectal & skin probe is secure and still in place at 2cm depth
- Address any alarms, check Criticool settings and check for water leaks on both ends of blue hoses. If leaking noted please refer to trouble shooting Appendix 3

If rectal temperature <36.5°C for 2 temperature cycles increase the cot temperature by 1°C every 15 minutes until normothemia achieved.

If rectal temperature >37.5°C for 2 temperature cycles, decrease the cot temperature by 1°C every 15 minutes.

If after decreasing cot temperature by 2°C (2 temperature cycles) baby remains >37.5, put the machine into standby. Turn back on when baby 37°C.

**When temp between 36.5-37.5ºC**

- Check lines and ventilation, Curewrap secured and assign team members roles to prepare for transfer
- Transfer baby into cot with baby in Curewrap and Neohelp and Bilinest/nest
- Place gel wedge onto Curewrap above head and bring hoses down either side of baby towards feet of cot

- Record **core and surface temperature every 15 mins** this will be “1 temperature cycle”.
- Fill out Hotwheels Criticool data entry form

**Setting up Criticool Device**

- Fill Criticool water tank with 1L sterile water and turn system on
- Insert temp probe ‘GREEN’ lead to ‘Surface’ port and ‘Grey’ lead to ‘Core’ port
- Attach metal blue water hoses (‘Water in’ on the right and ‘water out’ on the left) to Criticool
- Attach CureWrap to other end of hose and unclamp CureWrap white clamps
- Attach surface probe near clavicle (do not cover with wrap) and insert 7G rectal probe 2cm and secure to thigh, using Sil-flex tape to protect the skin and Grip-Lok to attach
- Select ‘Normothermia’ mode on Criticool and set **Target Temp to 37ºC**
- Place baby (still in Neohelp) into CureWrap, making sure the bilisoft is between baby and CureWrap. Wrap the CureWrap around the head and body, leaving a finger breadth between baby and wrap. Ensure baby is wearing a hat and nappy.
- Position the baby in the bilinest. Please see the Criticool Mini laminated sheet for pictures

**If temperature <36.5**°**C**

**If temperature >37.5**°**C**

**On arrival**

- Plug cot to power source and check overhead warmer is turned on
- Check baby’s axilla temperature, remove any wet clothing/towels/linens. Ensure wool hat on baby
- Place baby in Neohelp^TM^ if <24 hours old, baby is requiring external humidification, or was born <27 weeks and is <2 weeks old (Diagram 2, Appendix 3)
- Overhead heater temperature can be adjusted while setting up the Criticool device

**Prior to arrival referring hospital**

- Plug cot to power source in ambulance/plane and **set cot temp 35ºC (or 33ºC if baby >5 days old)**
- Place all Criticool equipment, CureWrap, Neohelp, bilinest, bilisoft and linen inside the cot to prewarm
- If needed switch on Neopod humidifier to 37-39ºC + prime with sterile water just prior to arrival

REMEMBER TO FOLLOW STANDARD CARE UNTIL RANDOMISATION HAS OCCURRED
